# A Longitudinal Study of BNT162b2 Vaccine-Induced Humoral Response and Reactogenicity in Health Care Workers with Prior COVID-19 Disease

**DOI:** 10.1101/2021.03.18.21253845

**Authors:** Steven G. Kelsen, Alan S. Braverman, Mark O. Aksoy, Jacob A. Hayman, Puja Patel, Charu Rajput, Huaqing Zhao, Susan G. Fisher, Michael R. Ruggieri, Nina T. Gentile

## Abstract

**Background:** Current recommendations in the United States are that subjects with a previous history of COVID-19 disease receive the full 2 dose mRNA vaccine regimen. We tested the hypothesis that humoral immune responses and reactogenicity to a SARS-CoV-2 mRNA vaccine (BNT-162b2) differ qualitatively and quantitatively in subjects with prior SARS-CoV-2 infection versus infection-naïve subjects.

**Methods:** Health care workers (n=61) from a single academic institution with and without prior COVID-19 received two 30 µg doses of BNT162b2 vaccine 3 weeks apart. The COVID group (n=30) received vaccine approximately 7 months post infection. IgG antibody against the Spike receptor-binding domain (RBD), serum neutralizing activity and vaccine adverse reactions were assessed every 2 weeks for 56 days after the 1^st^ injection. A longitudinal design and long study duration allowed the onset, maximum response and initial decay rate of Spike IgG antibody to be assessed in each subject. In addition, Spike IgG antibody levels are expressed as µg / mL to provide normal values for clinical decision making.

**Findings:** Spike IgG responses were highly variable in both groups. However, the COVID group manifested rapid increases in Spike IgG antibody and serum neutralizing activity post 1^st^ vaccine dose but little or no increase in Spike IgG or serum neutralizing activity after the 2^nd^ dose. In fact, Spike IgG was maximum prior to the 2^nd^ dose in 36% of the COVID group and 0% of controls. Peak IgG antibody was lower but appeared to fall more slowly in the COVID than in the control group. Finally, adverse systemic reactions e.g., fever, headache and malaise, after both the 1^st^ and 2^nd^ injection were more frequent and lasted longer in the COVID group than in the control group.

**Conclusions:** Health care workers with prior COVID-19 demonstrate a robust, accelerated humoral immune response to the 1^st^ dose of the COVID-19 mRNA vaccine but attenuated response to the 2^nd^ dose. They also experience greater reactogenicity than controls. Accordingly, subjects with prior COVID-19 may require only a single dose of vaccine.

## INTRODUCTION

Vaccination against the SARS–CoV–2 virus affords a way to rapidly achieve widespread, protective immunity of the uninfected population and thus end the COVID-19 pandemic. In fact, phase 2/3 clinical trials of the “Pfizer”-BNT162b2 and “Moderna”-mRNA-1273 mRNA vaccines now in use, demonstrate 90-95% protection against SARS-CoV-2 infection and 100% effectiveness in preventing severe COVID-19 disease, e.g., hospitalization or death.^1-5^ Similar protective efficacy of the Pfizer BNT162b2 vaccine has been reported in a country-wide population study in Israel.^3^

Public health authorities in the US recommend full vaccination including both doses of the two dose regimen mRNA vaccines for all those over the age of 12 years including those with prior SARS-CoV-2 infection or COVID-19 disease.^6^ Interestingly, this recommendation has been made despite the fact that trials of both COVID-19 mRNA vaccines excluded volunteers with a history of COVID-19 disease and most subjects post COVID-19 have durable immune memory.^7-11^ Moreover, the rate of SARS-CoV-2 re-infection in the 15 months since the pandemic started has been quite low.^12-14^

Of interest in this regard, several studies have demonstrated that subjects with prior SARS-CoV-2 infection / COVID-19 disease exhibit rapid and robust humoral and cell-mediated immune responses to a single dose of a two dose mRNA vaccine regimen.^15-23^ Moreover, responses of those with prior SARS-CoV-2 infection to the 1^st^ dose greatly exceed the responses of infection-naïve subjects. This observation has led to the suggestion that a single dose of a COVID mRNA vaccine may be sufficient to provide adequate protection against infection for subjects with prior SARS-CoV-2 infection.^18,24^ Given the scarcity of vaccines in most of the world, it has been suggested that subjects with a history of prior SARS-CoV-2 infection receive only 1 dose of the vaccine.^18,20,22^ In this approach, the second dose would be withheld until some future date as needed.

In contrast to the considerable available data defining the response to the 1st dose of an mRNA vaccine of SARS-CoV-2 infected / COVID-19 disease subjects, little or no information is available regarding their response to the 2^nd^ dose of vaccine given at the standard 3 to 4 week dosing interval. Anecdotal data, however, suggest that after a 2nd dose of an mRNA vaccine at least some subjects with prior COVID-19 disease exhibit reductions in protective IgG antibody levels and circulating memory B and T cells.^19^

Accordingly, this longitudinal study examined the hypothesis that the time course and magnitude of the humoral immune response and reactogenicity induced by a full, two dose mRNA vaccination regimen differed quantitatively in subjects with prior COVID-19 disease versus infection – naïve subjects. Specifically, we examined the level of anti-Spike IgG antibody and serum neutralizing activity serially at fixed points in time i.e., 2 week intervals, for 56 days after the 1st and 35 days after the 2^nd^ injection in SARS-CoV-2 infected and infection – naïve health care workers from the same academic health care center. To avoid possible confounding effects of differences in immune potency and reactogenicity between mRNA vaccines, a single vaccine i.e., Pfizer BNT162b2, was used.^25^

Our results indicate that Spike IgG antibody levels and serum neutralizing activity in response to BNT162b2 vaccine is time-dependent and that subjects with prior COVID-19 increase more rapidly, but reach lower peak levels and appear to fall more slowly than in infection-naïve subjects. Our data also represent the first available set of time-dependent normal values for Spike IgG antibody induced by BNT162b2 in normal adults with and without prior COVID-19.

## METHODS

Subjects recruited (n=61) into this vaccine study represented a subset of a larger cohort of healthcare workers (i.e., physicians, nurses, respiratory therapists or other ancillary health care personnel; n=281) participating in a surveillance study of SARS-CoV-2 Spike IgG seropositivity in our multi-hospital Health System. Subjects in the vaccine sub-study agreed to have blood samples drawn at 2-week intervals for 56 days following the initial dose of the BNT162b2 mRNA vaccine. Subjects completed a questionnaire detailing their history of SARS-CoV-2 infection / COVID-19 disease, job description, demographics and comorbidities. The study was approved by our Institutional Review Board.

Subjects in the SARS-CoV-2 infection / COVID-19 positive group (COVID group; n = 30) had a documented history of COVID-19 with a positive nasopharyngeal swab for virus RNA or were seropositive for IgG antibody against the Spike or nucleocapsid proteins. Subjects in the SARS-CoV-2 infection naïve group (control group; n=31) did not have: COVID-19 illness; viral RNA detected by PCR testing; and were IgG seronegative for the Spike and nucleocapsid proteins.

The BNT162b2 vaccine was given to all subjects as currently recommended i.e., two, 30 µg, 0.5 ml intramuscular injections given 3-weeks apart. Vaccine administration in both groups took place from December 16, 2020 until April 2, 2021. Blood samples were obtained at 14, 28, 42, and 56 days post 1^st^ dose. Those sampling intervals were based on the BNT162b2 phase 1 trial which demonstrated maximal immune responses by day 42.^5^

### SARS-CoV-2 Antibody

Serum SARS-CoV-2 Spike receptor binding domain (RBD) IgG antibodies were quantified by a two-step immunoassay using the Beckman Coulter, Access® microparticle-based system run on a high throughput UniCel Dxl 800 device (https://www.beckmancoulter.com/en/products/immunoassay/access-sars-cov-2-igg-ii-assay). This assay uses antigen-coated paramagnetic particles which when mixed with subject serum create an antigen-antibody complex. Anti-human IgG acridinium-labeled conjugate is then added to create a chemiluminescent signal measured as relative light units (RLU). Serum samples were diluted 1:10 and 1:20 to ensure that signals remained in the linear range of the standard curve. This system has a 4 order of magnitude dynamic range.^26,27^

A recent study by Bartsch et al indicates that Spike RBD IgG antibody concentrations when present above a critical threshold, correlate with other aspects of immune function such as serum neutralization activity, opsonization activity and T-cell activation responses to SARS-CoV-2 virus antigen.^28^ As such, the concentration of Spike IgG antibody seems to reflect the overall activity of the immune system. Accordingly, we used a recombinant, human IgG1 monoclonal SARS-CoV-2 Spike RBD antibody, Clone CR3022, produced in Nicotiana benthamiana (BEI Resources, Manassas, VA) to convert RLU values to µg/mL protein as previously described. ^28,29^

SARS-CoV-2 nucleocapsid IgG antibody was also measured to detect possible superimposed SARS-CoV-2 infection during the 56 days post vaccination. Nucleocapsid IgG antibody was measured using the Abbott Alinity system which is also a twostep, microparticle, chemiluminescent immunoassay (https://www.corelaboratory.abbott/us/en/offerings/segments/infectious-disease/sars-cov-2).

### Neutralization Assay

Serum neutralization assays were performed as previously described using luciferase – expressing, lentiviral particles pseudotyped for the SARS-CoV-2 Spike protein and HEK-293T cells overexpressing the ACE2 receptor. ^30,31^ The Spike protein was from SARS-CoV-2 strain Wuhan-Hu-1 as previously described by Crawford et al (see Supplementary Methods and Figures S3 and S4 for further details).^30^ Neutralizing activity was expressed as the serum dilution which produced 50% inhibition (IC_50_) of pseudo-particle entry. IC^50^ was calculated using sigmoidal 4 factor polynomial, non-linear regression in GraphPad Prism version 9.

### Vaccine Reactogenicity

The duration and severity of local and systemic reactions to the vaccine which occurred within 7 days of each injection were assessed using a standard questionnaire administered at the time of each blood draw. The severity of solicited local (pain / tenderness) and systemic reactions (fever, malaise, headache) were scored from 0 (none) to 10 (most severe) with scores ≥6 classified as severe.

### Statistical Analysis

Continuous measures within groups were expressed as mean ± 1 standard error (SE). Comparison of categorical variables between groups was assessed by Fisher Exact test and within groups by McNemar’s Test for paired comparisons. Comparison of changes in antibody levels over time, or between the COVID and control groups was assessed by linear mixed-effects models for repeated measures. Statistical significance of differences was accepted at the p<0.05 level.

## RESULTS

### Study Population

Demographic characteristics of COVID (n =30) and control groups (n= 31) are shown in **Table 1**. The two groups were well matched for age, gender and ethnicity. Mean age in the COVID and control groups was 47 years and 45 years, respectively (p>0.80). Gender was male in 50% of the COVID and 52% of the control group (p>0.80). Most subjects were Caucasians (80% and 77%, in the COVID and control groups, respectively, p>0.90) but African-Americans and Asians were also included in both groups.

**Table 1.** Subject Demographics.

|  | **COVID**<br>**(n=30)** | **Control**<br>**(n=31)** | **p**<br>**value** |
| --- | --- | --- | --- |
| **Age (years)**<br>Mean ± SEM | 47±3 | 45±2 | 0.82 |
| **Gender % (n)**<br>Males<br>Females | 50 (15)<br>50 (15) | 52 (16)<br>48 (15) | 0.85 |
| **Ethnicity % (n)**<br>Caucasian<br>Asian<br>African-American | 80 (24)<br>10 (3)<br>10 (3) | 77 (24)<br>13 (4)<br>10 (3) | 0.94 |

Clinical features of the COVID illness are shown in **Table 2**. Most subjects (93%) were symptomatic; 7% were asymptomatic. Four subjects (13%) had COVID–pneumonia. Two subjects (7%) were hospitalized.

**Table 2.**
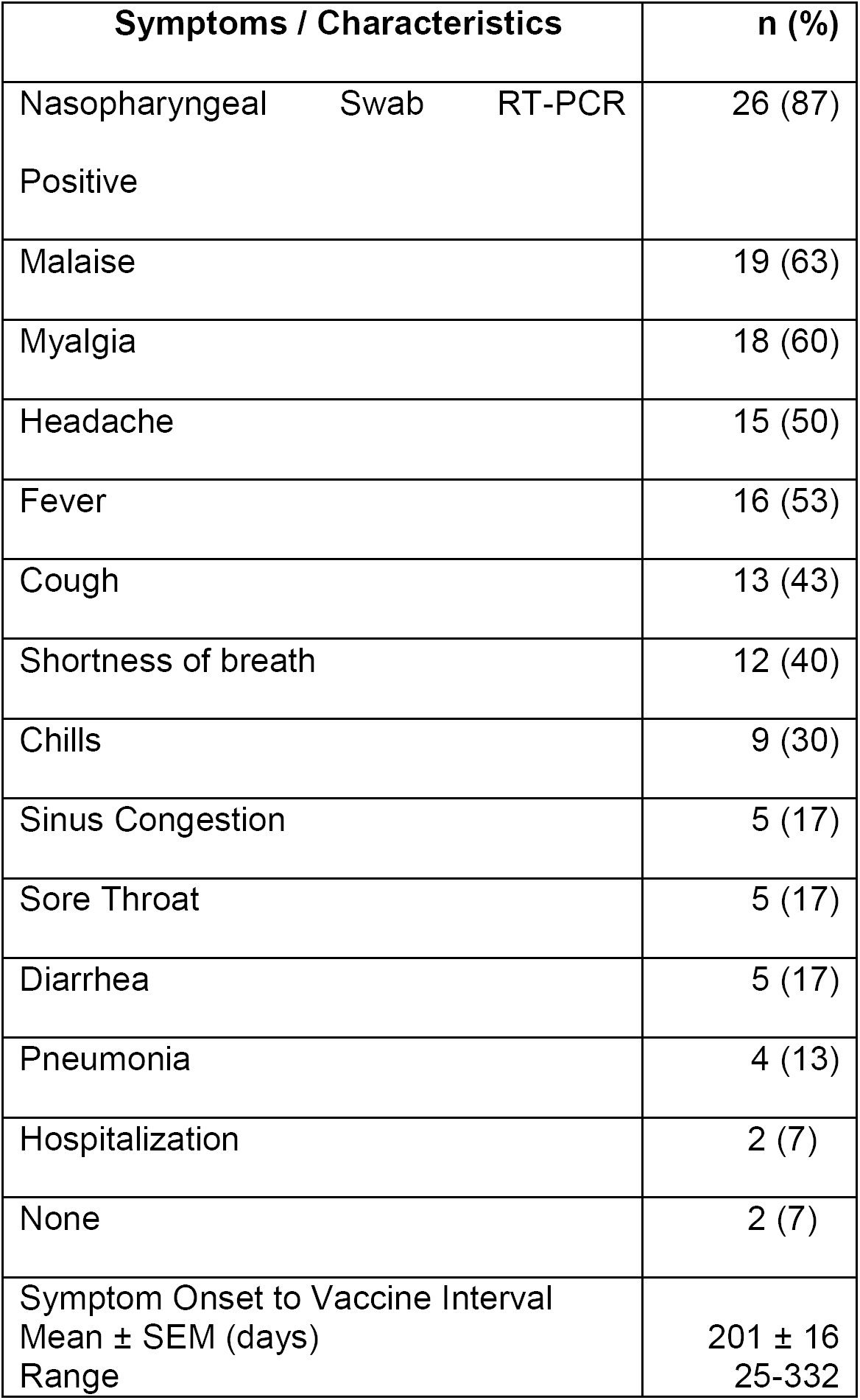
SARS–CoV-2 Infection (n=30)

Pre-vaccination levels of Spike RBD IgG or nucleocapsid IgG antibodies were elevated in 81% of subjects in the COVID group and none of the controls (**Figure S1**).

On average, COVID subjects received the first dose of vaccine ∼ 7 months post onset of symptoms (POS) (mean 201 ± 16 SE days; range = 25 to 332 days) (**Table 2**). In fact, 58% of the COVID group received the first vaccine dose more than 7 months POS (i.e., >220 days).

### Antibody Responses to Vaccine

Spike RBD antibody levels at each time point post vaccination are shown in **Figure 1 upper and lower panels** as arbitrary units and in **Table 3** as mcg protein / mL.

**Table 3.**
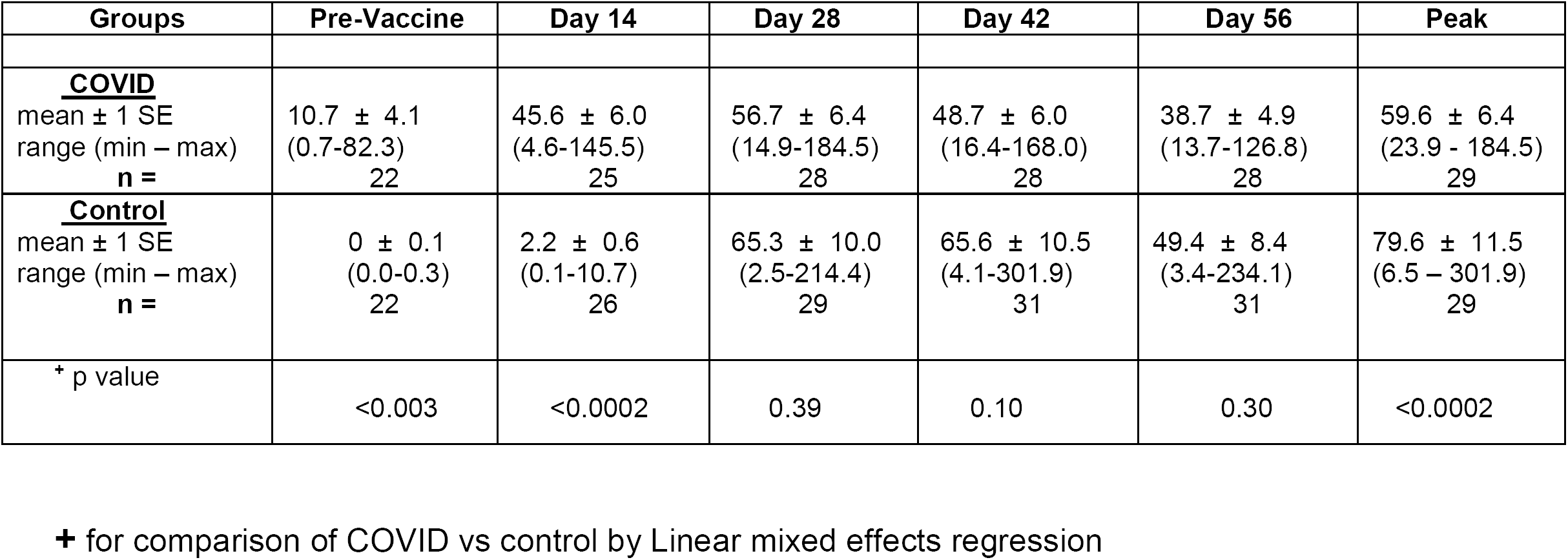
Spike RBD IgG Antibody Levels (mcg protein / mL serum)

**Figure 1.**
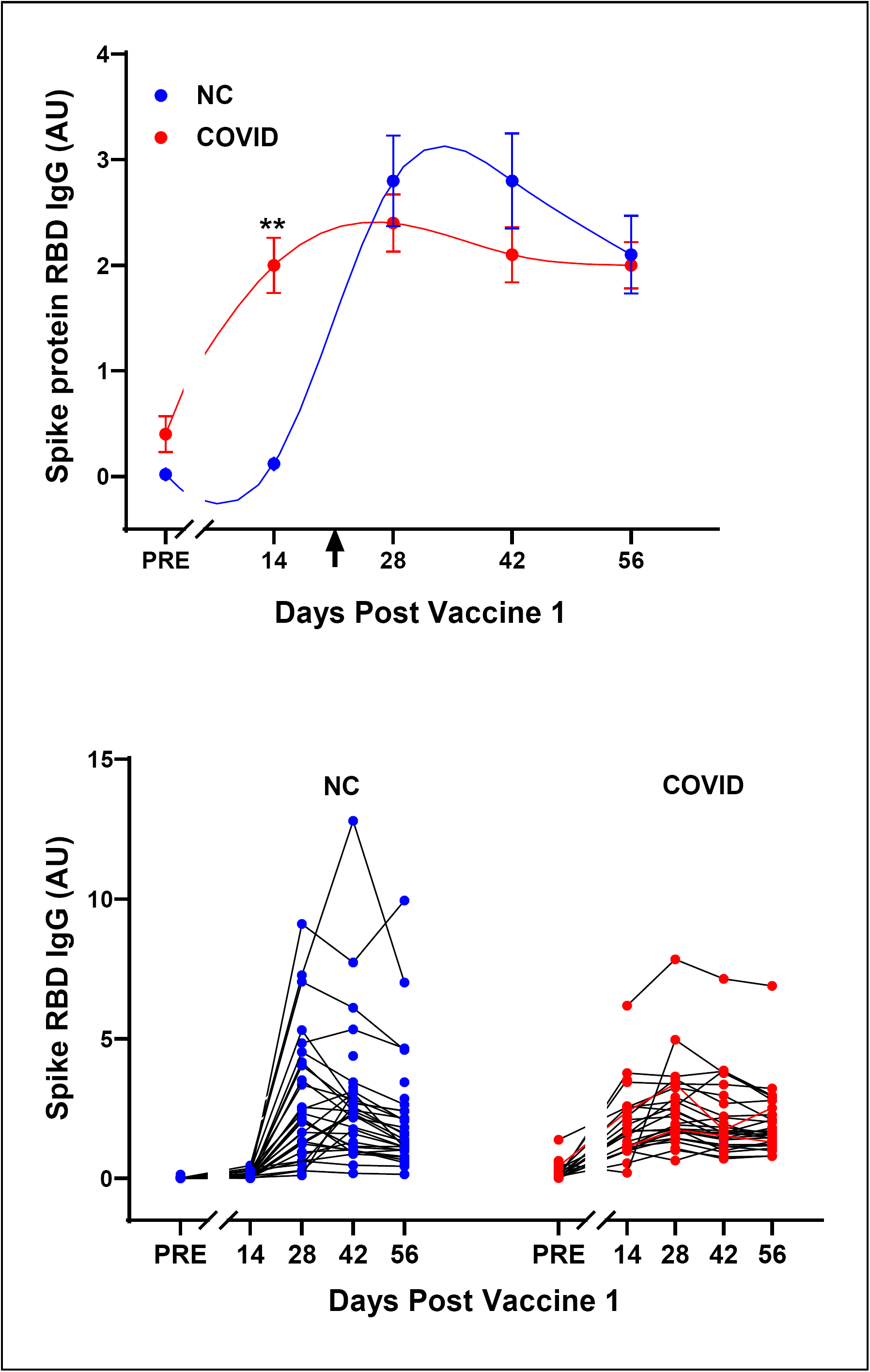
**Spike RBD IgG antibody responses to the BNT162b2 vaccine in COVID 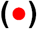 and control 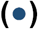 groups.** **Upper Panel:** Shows group mean ± 1SE responses. Vertical arrow indicates time of second vaccine injection. Note that the time course of Spike RBD IgG antibody response to vaccination was significantly different between COVID and control groups (p<0.0001; linear mixed effects model for repeated measures). Differences in Spike IgG antibody levels were significant pre-vaccine (p<0.003) and at day 14 (**p<0.0002). However, Spike IgG levels were similar at days 28, 42 and 56 (p>0.10 for all 3 comparisons). Sample size in the COVID group was: day 14 (n=25), day 28 (n=28), day 42 (n=28) and day 56 (n=28); in the control group: day 14 (n=26), day 28 (n=29), day 42 (n=31) and day 56 (n=31). **Lower Panel:** Spike RBD IgG antibody responses to vaccine in individual COVID and control subjects. Note the considerable inter-subject variability in both groups.

The time course and magnitude of the Spike RBD antibody response differed greatly across individuals in both groups (**Figure 1 lower**) but were significantly different in the COVID and control groups as a whole (p<0.0001 by linear mixed effects; **Figure 1 upper**). In the COVID group, Spike RBD IgG increased more rapidly, peaked earlier but appeared to fall more slowly than in the control group.

Specifically, in the COVID group, Spike IgG antibody at day 14 post 1^st^ injection increased significantly from the pre-vaccine level (p<0.0001) **(Table 3)**. Thereafter, little or no increase in Spike IgG occurred in the COVID group and IgG values at subsequent time points were not significantly different from the day 14 value (p>0.05 for all later time points compared with day 14). In the COVID group, Spike IgG levels peaked at day 14 in 36%; at day 28 in 52%; at day 42 in 12%; and at day 56 in 0% of subjects (**Figure 1 lower panel)**.

In contrast, in the control group, increases in Spike IgG at day 14 were not statistically different from the pre-vaccine value (p>0.20) and levels were significantly less than in the COVID group (p<0.0002). Thereafter, Spike IgG levels increased markedly between day 14 and day 28 in the control group (p<0.0001 for comparison of the 2 time points). In the control group, Spike IgG peaked at day 14 in 0%; at day 28 in 60%; at day 42 in 37%; and at day 56 in 3% of subjects. Of note, peak Spike IgG values were significantly greater in the control than the COVID group (p< 0.0002) **(Table 3**).

Spike IgG antibody levels declined from peak values in both groups but appeared to do so more slowly in the COVID group. In particular, from day 42 to day 56, Spike IgG fell significantly in the control group (p<0.001) but not in the COVID group (p>0.90).

The considerable variation in the interval between SARS-CoV-2 illness and vaccination i.e., post onset symptoms (POS), in the COVID group, i.e. 25 to 332 days (**Table 2**) did not appear to explain differences in peak Spike IgG levels. In fact, the peak Spike IgG level was unrelated to the interval between POS and vaccination (**Figure 2;** R^2^ = 0.01 by linear regression).

**Figure 2.**
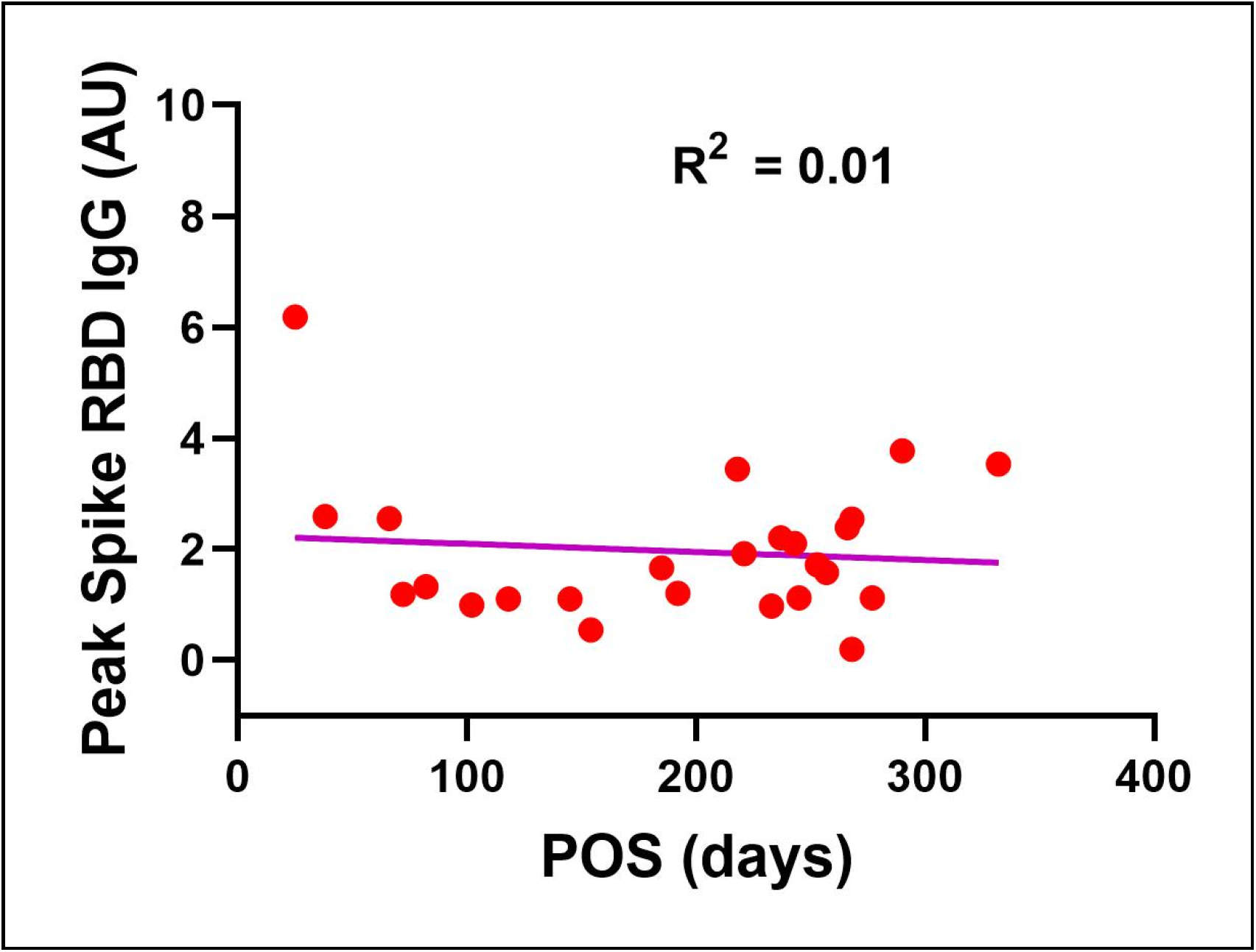
Relationship between vaccine-induced peak Spike RBD IgG antibody and interval post onset SARS-CoV-2 symptoms (POS) in COVID subjects (n=27). There was no discernable relationship (R^2^=0.01 by linear regression). Of note, 2 of the 29 subjects in the COVID group were asymptomatic. Hence, no POS value is available.

In contrast to Spike IgG, nucleocapsid IgG antibody, which was assessed to detect the unlikely possibility of superimposed SARS-CoV-2 infection during the study, was undetectable in the control group and declined progressively in the COVID group (**Figure S2**). Individual nucleocapsid IgG values are also shown in **Figure S2 (lower panel)**.

### Serum Neutralization Activity

Pre-vaccination, serum neutralization activity was modestly elevated in the COVID versus control group (IC_50_ 7×10^−3^ dilution and 2 × 10^−2^ dilution, respectively) but was not statistically significantly different in the 2 groups (p=0.20 by two way ANOVA) (**Figure 3 upper panel**).

**Figure 3.**
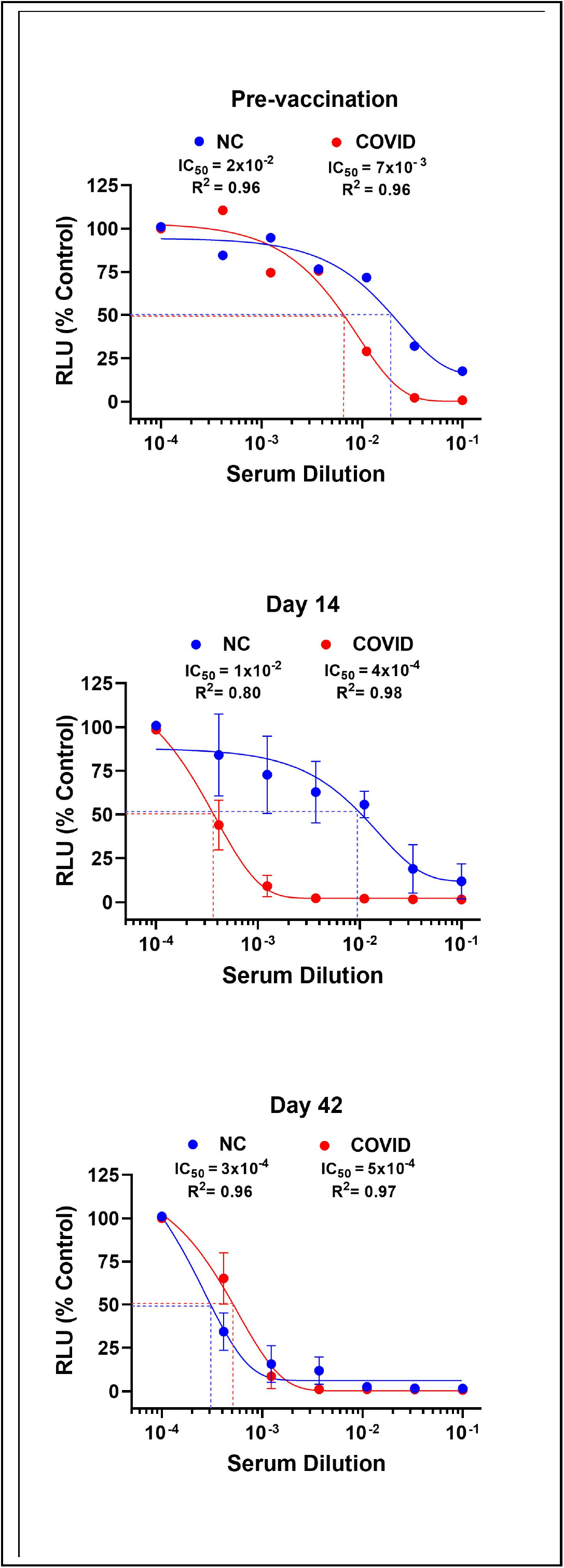
**Serum neutralizing activity in COVID 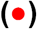 and control 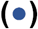 groups.** Pseudovirus uptake in HEK-293 ACE2 overexpressing cells was assessed from luciferase activity, i.e., relative light units(RLU), on the Y axis. Serum dilution is on the X-axis. The 100% control value on the Y axis represents maximal virus uptake occurring in the absence of serum. IC_50_ was calculated using sigmoidal 4 factor polynomial, non-linear regression Data are shown pre-vaccination **(upper)** and day 14 **(middle)** and day 42 **(lowe**r **panel)** post 1^st^ vaccine dose. Pre-vaccine, neutralizing activity in the COVID group (n=7) tended to be greater than in the control group (n=7) (IC_50_ 7 × 10^−3^ vs 2 × 10^−2^ dilution, respectively) but was not statistically significantly different (p=0.20 by 2-way ANOVA). At Day 14 post 1^st^ injection, neutralizing activity increased greatly in the COVID group (n=21) but was unchanged in the control group (n=21) (IC_50_ = 4 × 10^−4^ vs 1 × 10^−2^ dilution, respectively; p < 0.03 by 2-way ANOVA for comparison of the 2 groups). In contrast, at Day 42, neutralizing activity increased greatly in the control group (n=21) but only slightly in the COVID group (n=21) (IC_50_ = 3 × 10^−4^ vs 5 × 10^−4^ dilution, respectively) and was again not statistically significantly different (p = 0.11 for comparison of the 2 groups by 2-way ANOVA). For days 14 and 42, the same 21 COVID and 21 control subjects were studied, and for both groups each datapoint is the mean ± SE for 3 pools of 7 subjects each.

At day 14 post 1^st^ injection, neutralizing activity increased ∼17 fold in the COVID group but was unchanged in the control group **(Figure 3 middle panel)**. Specifically, IC_50_ was 4 × 10^−4^ dilution in the COVID group and 1 × 10^−2^ dilution in controls i.e., ∼25 fold greater in the COVID group (p<0.03 by two way ANOVA).

At day 42 post 1^st^ injection, however, neutralizing activity in the COVID group was essentially unchanged from the 14 day value (i.e., IC_50_ 4 × 10^−4^ dilution and 5 × 10^−4^ dilution for days 14 and 42, respectively). In contrast, neutralizing activity increased markedly in the control group from the day 14 value (i.e., IC_50_ 1 × 10^−2^ dilution and 3 × 10^−4^ dilution for days 14 and 42, respectively) **(Figure 3 lower panel)**. As a result, there were no differences in neutralizing activity in the COVID and control groups at day 42 (p=0.11 by two way ANOVA for the 2 curves).

### Vaccine Reactogenicity

In general, systemic reactogenicity was greater in the COVID than the control group. Specifically, after the first injection, systemic symptoms (i.e., fever, headache, malaise/fatigue) were more frequent (p<0.05 for each by Fisher’s exact test) and lasted longer (p<0.001 by unpaired t test) in COVID than in control subjects **(Fig. 4A and B)**. The use of antipyretics and/or analgesics was also more frequent in COVID (p<0.05 by Fisher Exact test) **(Fig. 4B)**.

**Figure 4:**
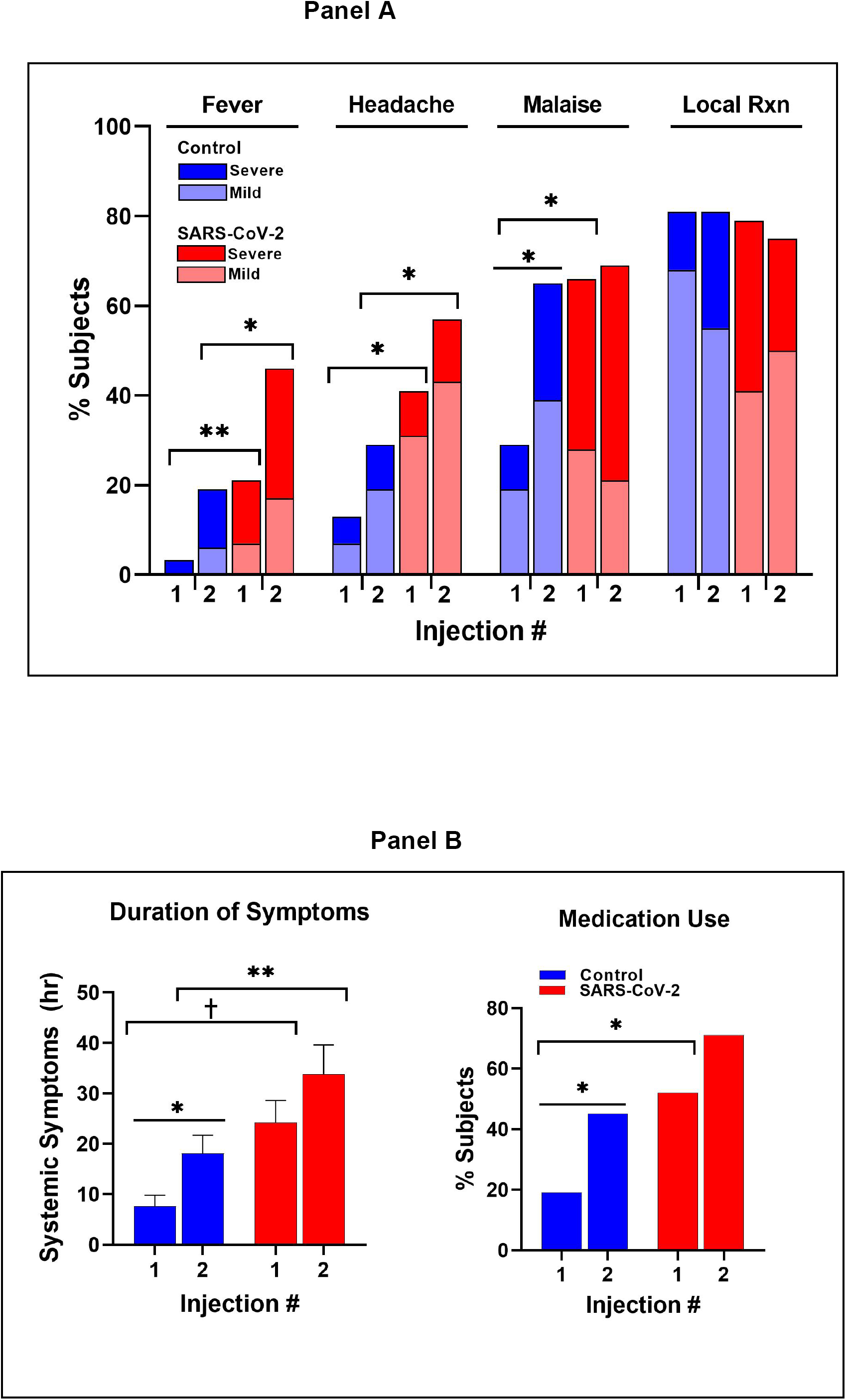
Reactions to the 1^st^ and 2nd Pfizer BNT162b2 mRNA vaccine in COVID and control groups. **Panel A:** Prevalence and severity of systemic and local reactions. Dark color (red or blue) indicates reaction scores of > 6 severity; light colors indicate scores ≤ 5. Brackets indicate statistical comparisons across groups. Lines indicate comparisons within groups. Asterisk (**\***) indicates p<0·05; double asterisk (**\*\***) indicates p<0·01; and cross (**†**) indicates p<0·001. **Panel B:** Duration of systemic symptoms (**Left**) and frequency of medication use **(Right)** in COVID and control groups.

In contrast, local reactions (i.e., pain and tenderness) occurred in most subjects (≥80%) and with similar frequency in both groups (p=NS) (**Fig. 4A)**.

After the second injection, fever and headache were again more frequent in the COVID group (p<0.05 for both) and systemic symptoms continued to last longer (p<0.01) **(Fig. 4A and B**). Local reactions to the 2^nd^ injection were similar in frequency and severity in both groups **(Fig. 4A)**.

All reactions resolved within 7 days without need for medical attention.

## DISCUSSION

This study in a cohort of healthcare workers in a single academic medical center was designed to test the hypothesis that humoral immune responses and reactogenicity to a SARS-CoV-2 mRNA vaccine (BNT-162b2) differ in subjects with and without prior SARS-CoV-2 infection. This hypothesis was based on observations that subjects with prior SARS-CoV-2 infection have long-lasting memory B-cell and T-cell based immunity to the Spike protein immunogen in the vaccine.^7,10^ Accordingly, to define the vaccine-induced, humoral immune response and reactogenicity, subjects were studied at 2-week intervals for 56 days post initial vaccination. The longitudinal design and length of the study allowed the onset, maximum response and initial decay rate of Spike IgG antibody to be assessed in each subject.

Our results indicate that the time course and magnitude of the Spike RBD IgG and serum neutralizing responses to the vaccine differed in the two groups. Specifically, in the COVID group, Spike IgG antibody increased more rapidly, but reached lower peak levels and seemed to fall more slowly i.e., the response was “flatter,” in the COVID group than in the infection-naïve, control group. In fact, a large percentage of the COVID group (36%) achieved maximum Spike IgG antibody responses 14 days after the initial injection and did not respond to the 2^nd^ injection. Moreover, for the COVID group as a whole, serum neutralizing activity was maximum at day 14 and did not increase with the 2nd vaccine. Accordingly, serum neutralizing activity mirrored the Spike IgG antibody response.

In contrast to changes in Spike IgG, nucleocapsid IgG was undetectable in the control group and decreased progressively over the observation period in the COVID group. This finding rules out the remote possibility that superimposed SARS-CoV-2 infection in either group during the vaccination period may have confounded the results.

The frequency and duration of systemic reactions to the BNT-162b2 vaccine were heightened in subjects with prior SARS-CoV-2-infection although none were serious. Heightened systemic reactions in the COVID group were present with both the 1^st^ and 2^nd^ injection but were most apparent after the 1st injection.

The rapid, robust Spike IgG response to the first dose of vaccine in the COVID group more than 7 months after prior infection and attenuated response to the second dose likely represents an anamnestic response mediated by long duration memory B and T cells.^7,10,17^ In fact, most SARS-CoV-2 infected individuals continue to have circulating memory B-cells, bone marrow plasma cells, T follicular helper cells and CD4 and CD8 Th1-cells present for 8 to 11 months post infection.^7,8,10^ Pre-existing immune memory against the Spike immunogen in the COVID group may also explain the heightened vaccine reactogenicity.

Our study has a number of strengths. First, the longitudinal study design with fixed 2-week sampling intervals and long duration allowed us to define the onset, peak and initial decay rate of Spike IgG antibody in both groups despite the considerable individual variability.

Second, the COVID and control groups were well-matched for age and gender. This is important since age and gender determine immune responses to many vaccines.^32,33^ Accordingly, differences in vaccine responses observed in the 2 groups in this study are not explainable by differences in gender or age.

Third, accepted methods were utilized to assess Spike RBD IgG antibody levels.^26,27,34^ In fact, antibody levels were expressed in absolute units (i.e., mcg protein / mL) as well arbitrary units (i.e., chemiluminescence units).^28,29^ We specifically chose to express the Spike IgG response as mcg protein / mL to allow the “normal” vaccine response of healthy subjects to be available for medical decision making. That is, Spike IgG levels generated in a healthy population at discrete points in time post-vaccination are now available to assess the adequacy of the vaccine response in subjects with co-morbid medical conditions which could impair the immune response (e.g., solid organ transplant).^35^

We assessed the serum neutralizing activity in the two groups using an accepted pseudotyped lentivirus neutralization assay since not all antibodies targeting the RBD are neutralizing.^30,36,37^ Conversely, antibodies against Spike protein epitopes outside the RBD may also be neutralizing.^36,37^ Accordingly, serum neutralizing activity represents a more comprehensive way of assessing the humoral immune response.^34^

Fourth, since both the immune response and adverse effects induced are vaccine-type dependent, a single, extensively used vaccine i.e., BNT162b2, was studied to avoid confounding effects on the responses observed.^25^

Our study also has limitations. First, while our data over 56 days post 1^st^ injection suggest that Spike IgG antibody levels may fall more slowly in the COVID group than in infection naïve subjects, the long-term IgG antibody level was not defined in this study. This is of importance since vaccine protection depends on the sustained antibody level.^32^ Additional time points will be needed in this regard.

Second, we studied subjects in the COVID group at a single time point i.e., ∼ 7 months post infection. Although no relationship between the interval post infection and peak Spike IgG antibody level was evident in the COVID group, it would be desirable to obtain data at additional intervals post infection. Also, our COVID group consisted almost entirely of symptomatic individuals (93%) biased toward the severe end of the spectrum (several had pneumonia and were hospitalized). Since the intensity of the humoral and cell-mediated responses to SARS-CoV-2 correlate directly with the number and severity of COVID-19 symptoms, subjects with milder forms of SARS-CoV-2 infection e.g., asymptomatic or pauci-symptomatic infection may respond differently.^19,38^

Third, our neutralizing assay utilized the original Wuhan-Hu-1 strain for the Spike protein as the neutralizing antibody target. Accordingly, our serum neutralizing activity study did not measure effectiveness against the more recent SARS-CoV-2 variants which are of considerable public health concern.

Finally, we did not assess other IgG antibody-mediated effects in addition to neutralization (e.g., opsonization, complement fixation and NK cell activation). However, in this regard, Bartsch at al observed in a cohort of subjects convalescing from SARS-CoV-2 infection that increasing levels of IgG Spike RBD antibody above a threshold of ∼0.5 – 1.0 mcg/mL indicate broad activation of the adaptive and innate immune system.^28^ Specifically, Bartsch observed that Spike RBD IgG antibody levels above this threshold correlate directly with increasing neutrophil phagocytosis, complement fixation and T-cell responses to SARS-CoV-2 Spike antigens. The maximal level of Spike IgG antibody in this cohort was 11 mcg / mL.^28^ In our study, the ∼45 mcg/mL Spike IgG antibody level in the COVID group at day 14 post vaccination suggests that broad immune activation e.g., Fcγ receptor-mediated activity and T-cell activation was likely achieved. Moreover, this degree of immune activity in the COVID group occurred before the second injection.

The results of our study are in agreement with recent studies of the immune response to the mRNA vaccines most of which were cross-sectional in design and focused on the response to the 1^st^ injection.^15-23^ Like the present study, they also report more rapid increases in Spike IgG antibody after the first injection of an mRNA vaccine in subjects with prior COVID-19 than in infection-naïve subjects. Our study extended these observations by more precisely defining the time course of the Spike antibody response, i.e., onset, peak and rate of decay to both doses of an mRNA vaccine. We also assessed serum neutralizing activity.

Moreover, our data represent the first available set of time-dependent “normal” values for Spike IgG antibody induced by BNT162b2 in healthy subjects with and without prior COVID-19. Accordingly, our data can be used to assess the level of the anti-Spike antibody response to BNT162b2 in potentially immunocompromised individuals e.g., solid organ transplant recipients.^35^ Our data, therefore, may facilitate medical decision making in the care of individual patients.

An important public health implication of our study is that subjects with a prior history of SARS-CoV-2 infection / COVID-19 may not respond to a second dose of an mRNA vaccine and hence may not need it. In essence, the prior bout of COVID-19 may have provided sufficient immune stimulation such that the first dose of vaccine elicited a maximal or near maximal response. In fact, anecdotal reports in small numbers of subjects with prior SARS-CoV-2 – infection indicate that the 2^nd^ injection may cause a reduction in Spike IgG levels and decreases in circulating B memory and T-cells.^19^

In conclusion, the present study indicates that humoral responses to an mRNA vaccine are time-dependent and differ in subjects with prior SARS-CoV-2 infection and infection – naïve subjects. Subjects with prior, generally moderate-severe COVID-19 disease achieve a rapid, maximal or near maximal level of humoral immunity after a single dose of a COVID mRNA vaccine. In fact, the humoral immune response to the second dose is greatly attenuated if not absent in subjects with prior COVID-19.

The possibility that a single dose of vaccine in subjects with prior COVID-19 is as efficacious as the 2 dose regimen in achieving a protective immune response has profound public health implications. It affords an opportunity to conserve millions of doses which could be used to help address the critical world-wide shortage of vaccine. This issue, however, will require a proper controlled trial in SARS-CoV-2-infected individuals in which the protection against re-infection with recent variants achieved with 1 vaccine dose is compared with the current 2 dose regimen.

## Data Availability

There are currently no external datasets or supplementary materials online at other repositories pertaining to this manuscript.

## ACKNOWLEDGEMENTS

The authors are grateful to the entire community of Temple University Hospital physicians, nurses, technicians and other health care workers who contributed considerable time over many months in support of this project. Their altruism and courage in the care of our patients with COVID-19 was a source of inspiration to us all.

We also wish to thank BEI Resources whose repeated, timely shipments of crucial reagents allowed us to carry out the assays on which this project was based.

The following reagents were obtained through BEI Resources, NIAID, NIH: Monoclonal Anti-SARS Coronavirus Recombinant Human IgG1, Clone CR3022 (produced in Nicotiana benthamiana), NR-52392; SARS-Related Coronavirus 2, Wuhan-Hu-1 Spike-Pseudotyped Lentiviral Kit, NR-52948; Human Embryonic Kidney Cells (HEK-293T) Expressing Human Angiotensin-Converting Enzyme 2, HEK-293T-hACE2 Cell Line, NR-52511; Monoclonal Anti-SARS-Related Coronavirus 2 Spike Glycoprotein RBD-mFc Fusion Protein (produced in vitro), NR-53795.

## SUPPLEMENT

### METHODS

#### Neutralization Assay

Serum neutralization assays were performed as previously described by Crawford et al using luciferase-expressing lentiviral particles pseudotyped for the SARS-CoV-2 spike protein and HEK-293T cells over-expressing the ACE2 receptor (HEK-293T-hACE2).^30^

##### Pseudovirus preparation

Pseudotyped lentivirus was generated in HEK-293T cells transfected simultaneously with the following: helper plasmids encoding for Gag and Pol, Tat1b, and Rev1b; lentiviral backbone plasmid encoding for Luc2 and ZsGreen; and plasmid encoding the Wuhan-Hu-1 strain of SARS-CoV-2 Spike glycoprotein (Lentiviral Kit, BEI Resources, #NR-52948). After 72 hrs, the culture supernatant containing pseudovirus was harvested, syringe-filtered (0.45 µm), and stored at -80° C.

##### Neutralization assay

HEK-293T-hACE2 cells (80,000 cells/well; BEI Resources, #NR-52511) were cultured in white-walled microplates for 24 hr. Pseudovirus entry was assessed from luciferase activity using a GloMax Discover luminometer (Promega Corp., #GM3000) and Bright-Glo reagent (Promega, #E2610). Initially, to determine optimum conditions for the assay, the signal generated by various pseudovirus dilutions was assessed in the absence of serum **(Fig. S3)**. A 10x-dilution of pseudovirus stock (plus 5 µg/mL polybrene, EMD Millipore Corp., #TR-2003-G) which yielded an RLU of 25,000 was used throughout. Negative controls i.e., supernatants from HEK-293T cells transfected with carrier DNA only (Promega, #E4881) (data not shown), or with a lentiviral plasmid lacking the Spike glycoprotein, yielded RLU values less than 0.2% of the undiluted pseudovirus (**Fig. S3**). For the neutralization assay itself, 7-point serial serum dilutions were incubated with pseudovirus for 1 hr at 37°C in a separate sterile plate. The serum-pseudovirus mixtures were then incubated with HEK-293T-hACE2 cells for 48 hr at 37° C. A potent neutralizing Spike RBD IgG antibody (BEI Resources, #NR-53795) was used as a positive control in each run (**Fig. S4**).

##### Experimental Design and Data Analysis

Neutralization assays were performed pre-vaccination and at days 14 and 42 post 1^st^ injection of vaccine. Individual serum samples (50 µL / subject) were pooled with 7 subjects per pool in both COVID and control groups. On days 14 and 42, data points for both groups are the mean ± SE of 3 pools of the same 21 COVID and 21 control subjects. In contrast, the pre-vaccination timepoint was a single pool for each group.

Maximal pseudovirus uptake by HEK cells in the absence of serum was taken as 100% of control. IC_50_ i.e., 50% inhibition of pseudovirus entry, was calculated using sigmoidal 4 factor polynomial, non-linear regression (GraphPad Prism version 9).

## FIGURE LEGENDS

**Figure S1.**
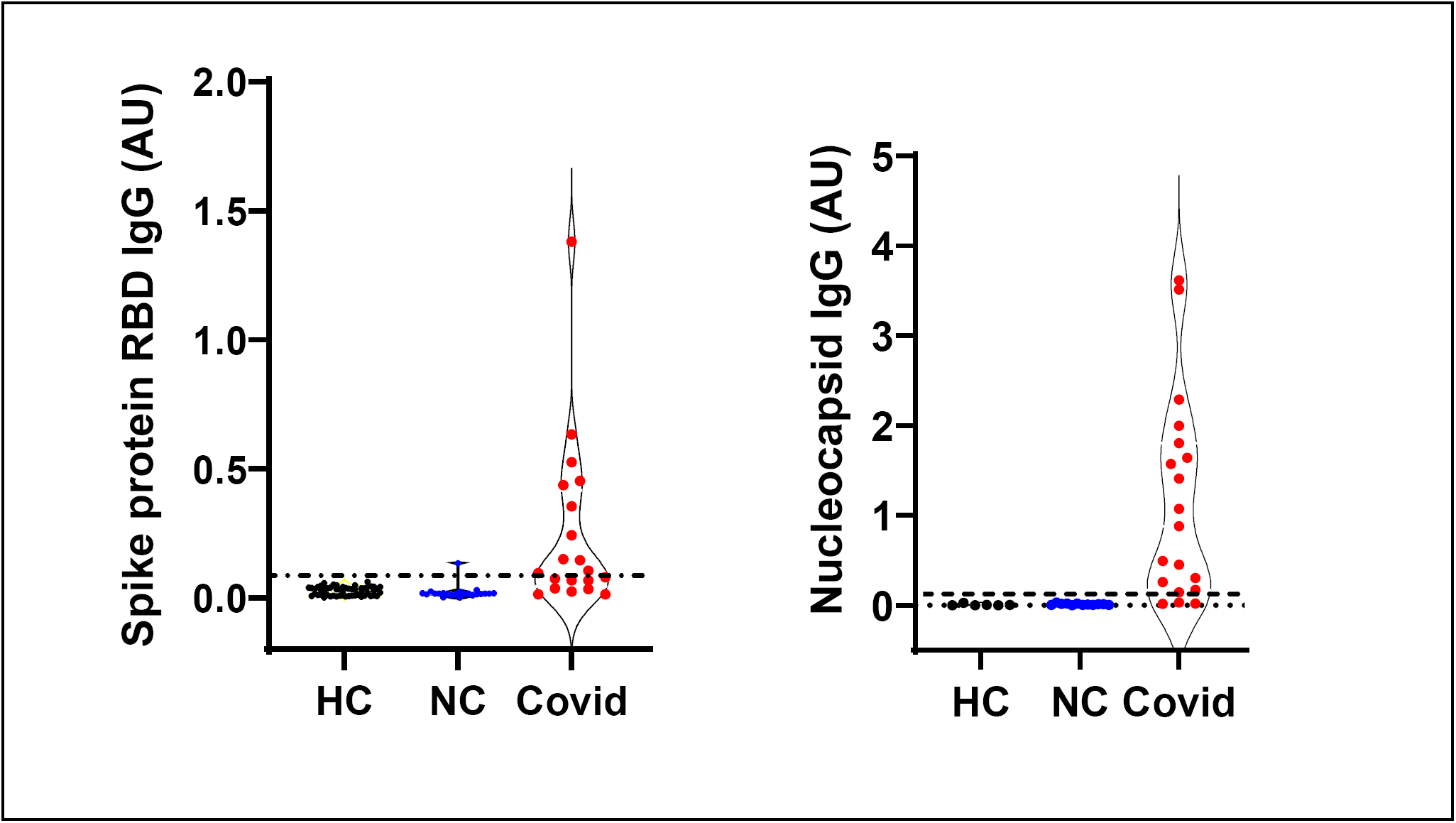
**Violin plots showing pre-vaccination serum Spike RBD IgG antibody (left) and nucleocapsid IgG antibody (right) in individual COVID (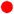, n=22) and control (NC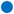, n=22) subjects.** Spike RBD IgG (p<0.01) and nucleocapsid IgG (p<0.003) antibody levels were significantly higher in the COVID compared to control groups prior to vaccination. Historical control samples (**HC**-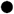; n=57) were archived prior to the COVID-19 pandemic i.e., 2008. Horizontal dashed line demarcates 4 standard deviations from the mean of the historical controls.

**Figure S2.**
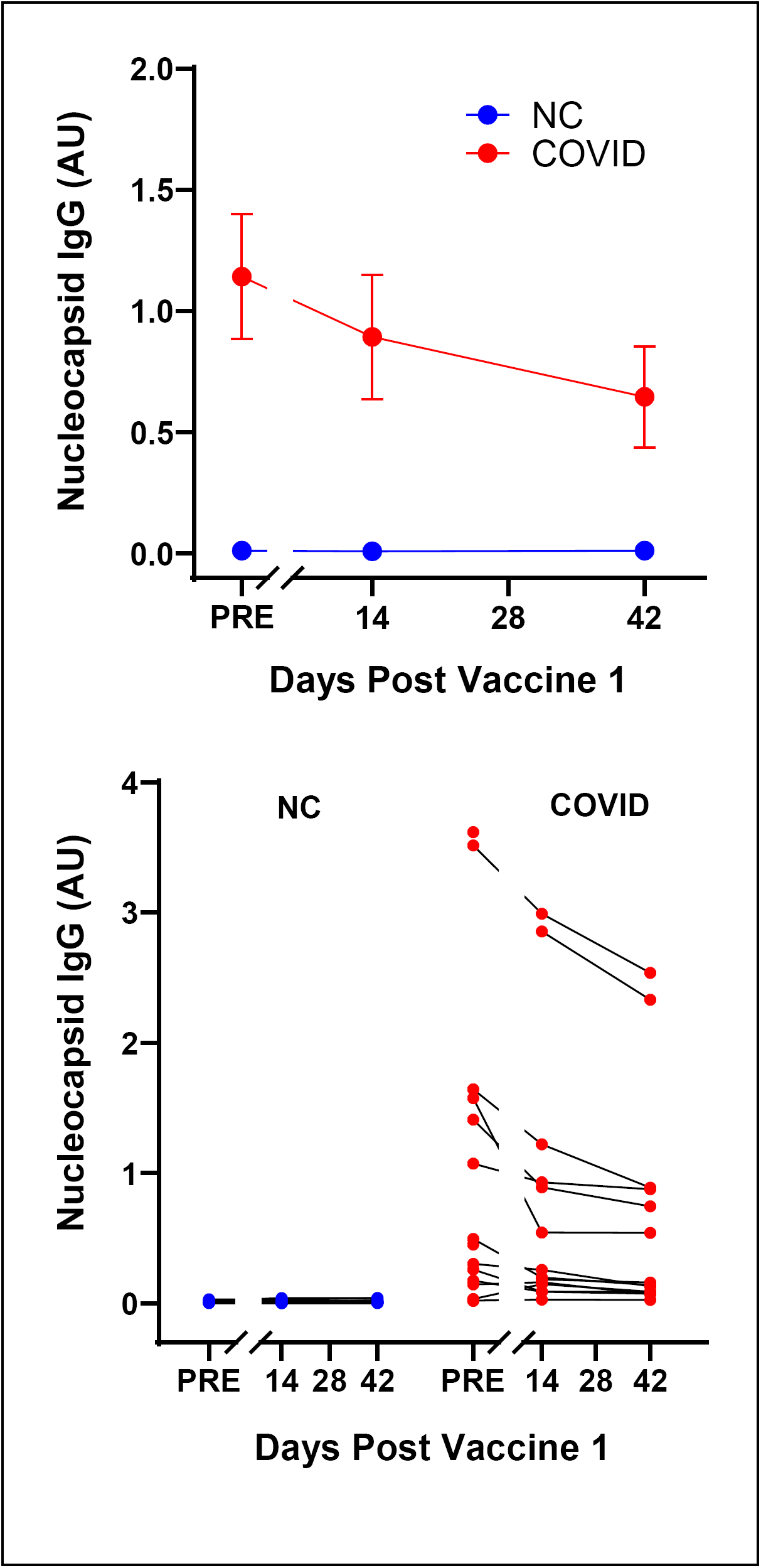
**Nucleocapsid IgG antibody in COVID 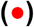 and control 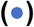 groups following the Pfizer BNT162b2 vaccine.** **Upper Panel:** Shows group mean ± 1 SE responses. Sample size for COVID group was: day 14 (n=16), day 42 (n=18). Sample size for control group was: day 14 (n=16), day 42 (n=17). **Lower Panel:** Individual COVID and control subjects. Note that nucleocapsid IgG antibody is undetectable in the control group and falls over time post vaccination in the COVID group.

**Figure S3.**
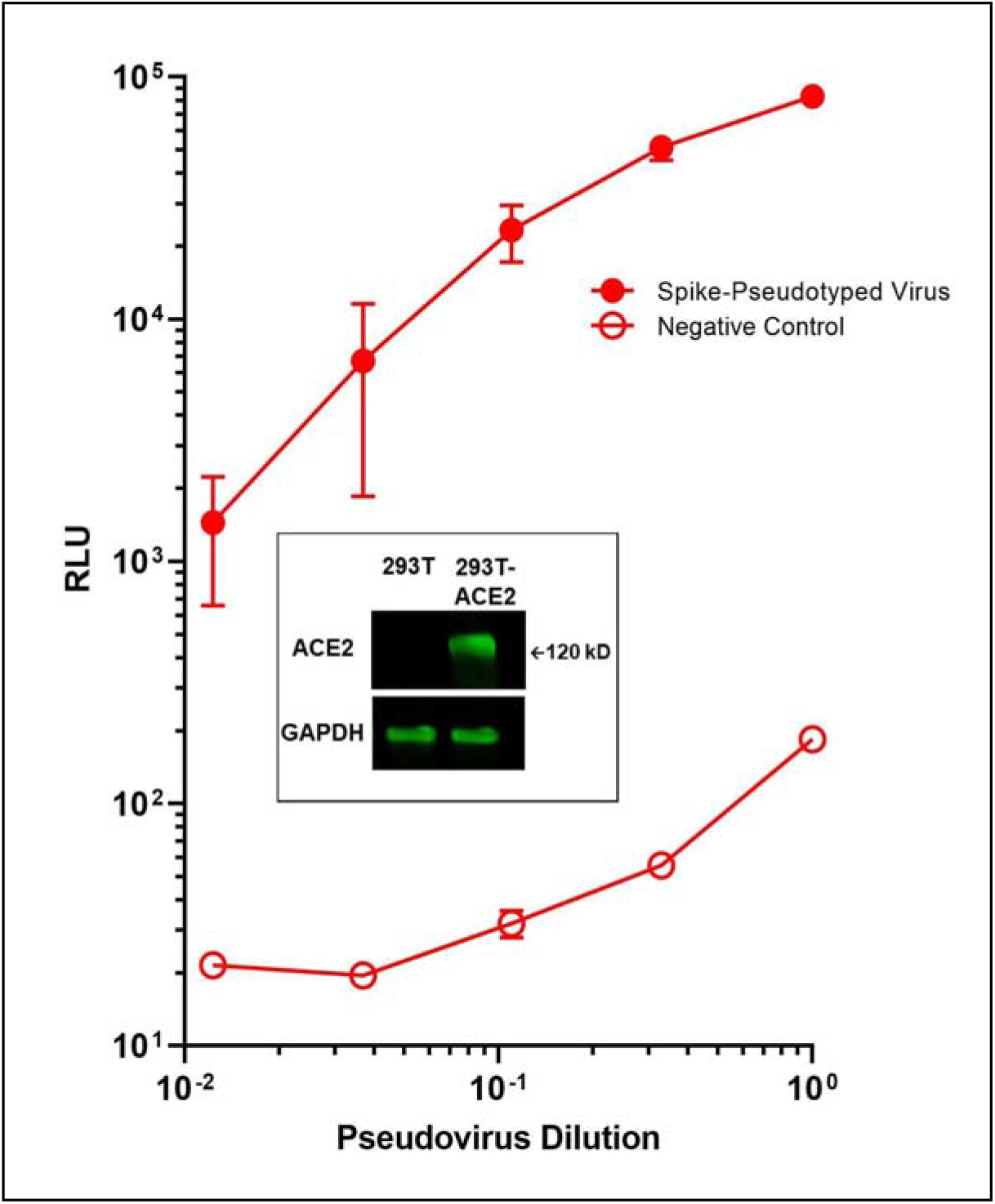
Uptake of pseudotyped SARS-CoV-2 lentivirus into HEK293T-ACE2 cells. Intact pseudotyped lentivirus 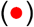 uptake shown as luciferase activity on the Y axis (relative light units - RLU) was assessed in the absence of serum. Pseudovirus dilutions are shown on the X axis. Lentivirus lacking SARS-CoV-2 Spike protein 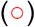 was used as an inactive, negative control. Data are Group mean ± 1 SE from multiple wells (1 experiment representative of 3). Note that intact pseudovirus diluted 10X from stock generated a signal 500 fold greater than that produced by the inactive pseudovirus. <u>Inset</u> - Western blot showing marked increase in ACE2 receptor expression in 293T-ACE2 vs native 293T cells.

**Figure S4.**
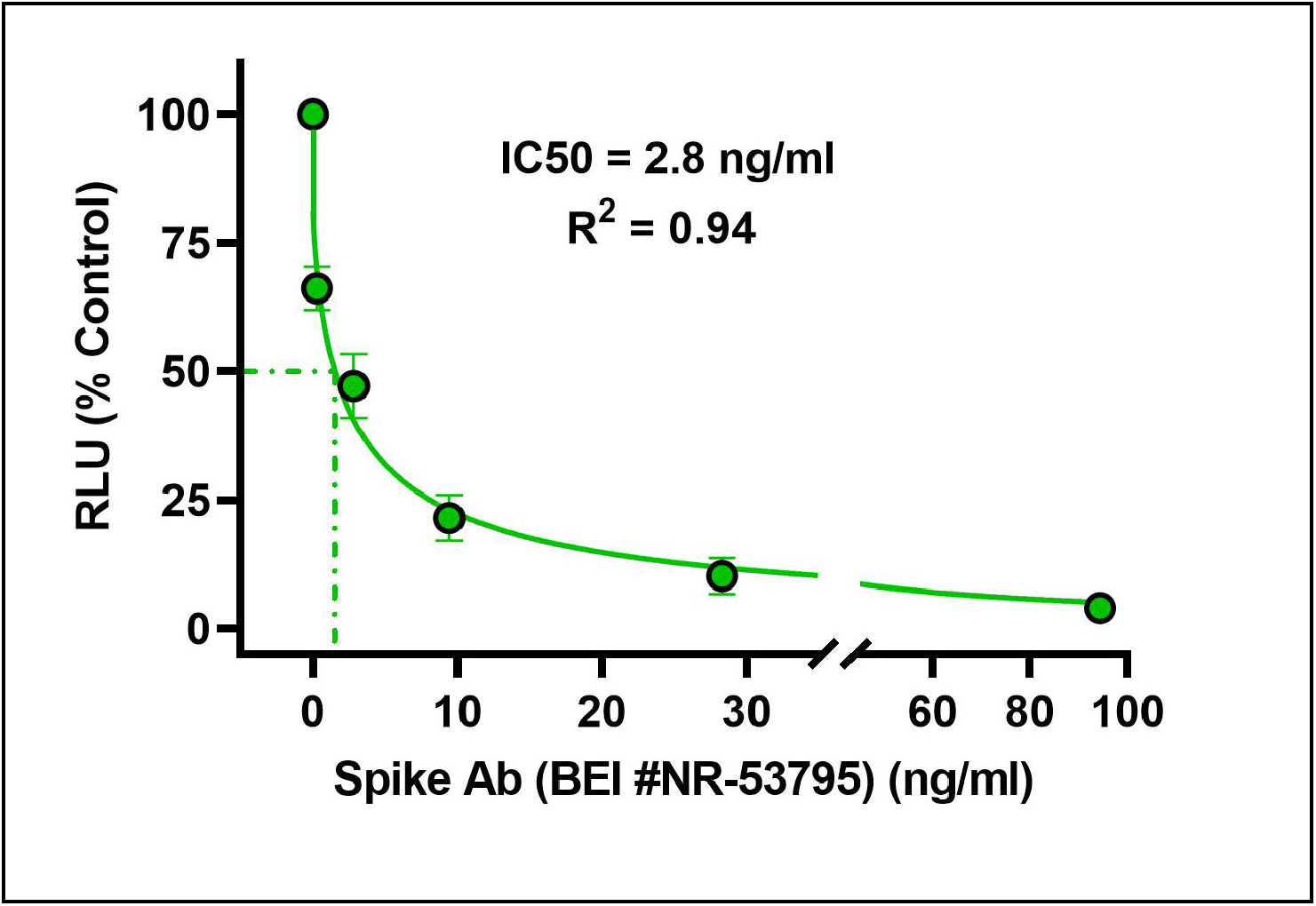
Effect of neutralizing Spike IgG antibody on pseudotyped SARS-CoV-2 lentivirus entry into HEK293T-ACE2 cells. Note that positive control Spike RBD neutralizing antibody (BEI #NR-53795) strongly inhibited pseudovirus uptake (IC_50_=2.8 ng/mL). Data are mean ± 1 SE of 4 experiments.

