## Supplementary Material for "A Longitudinal Study of BNT162b2 Vaccine-Induced Humoral Response and Reactogenicity in Health Care Workers with Prior COVID-19 Disease"

**Figure S4. Effect of neutralizing Spike IgG antibody on pseudotyped SARS-CoV-2 lentivirus entry into HEK293T-ACE2 cells.** Note that positive control Spike RBD neutralizing antibody (BEI #NR-53795) strongly inhibited pseudovirus uptake ($IC_{50}$=2.8 ng/mL). Data are mean ± 1 SE of 4 experiments.

Figure S1

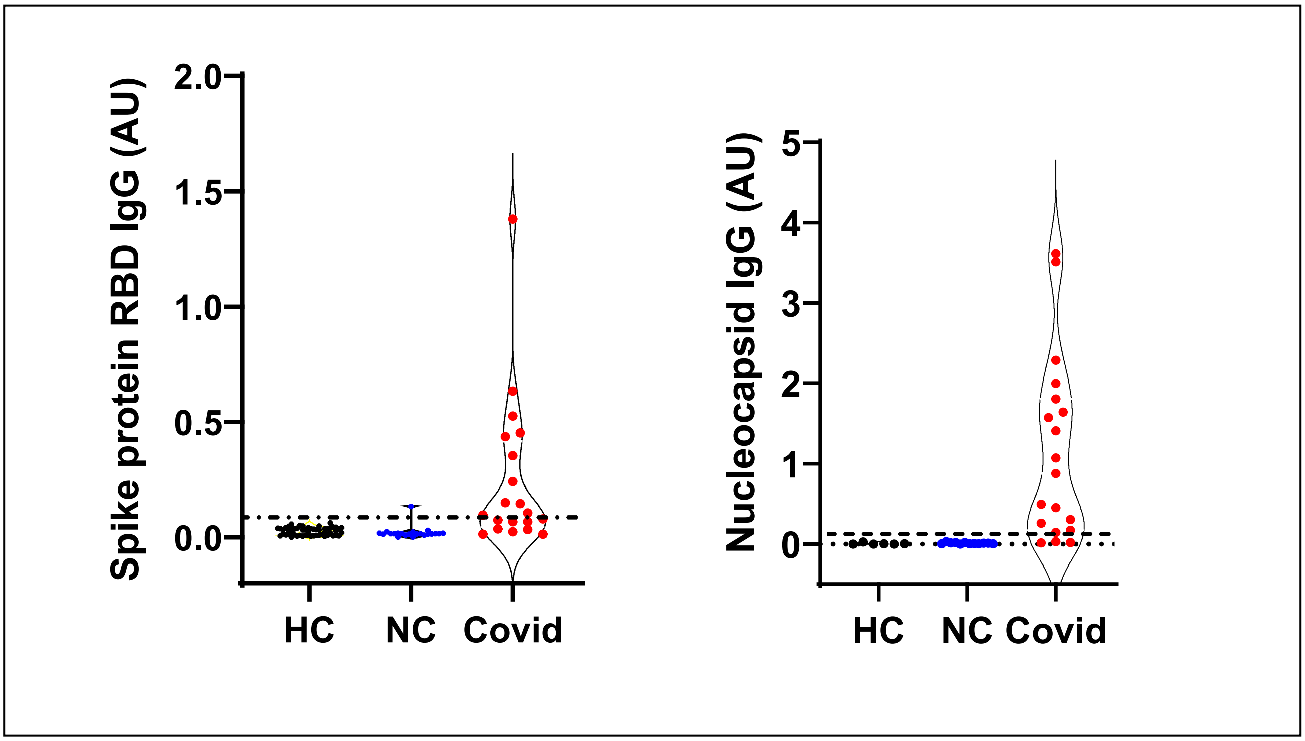
Spike protein RBD IgG (AU)
2.0
1.5
1.0
0.5
0.0
HC
NC
Covid
Nucleocapsid IgG (AU)
5
4
3
2
1
0
HC
NC
Covid

**Figure S2**

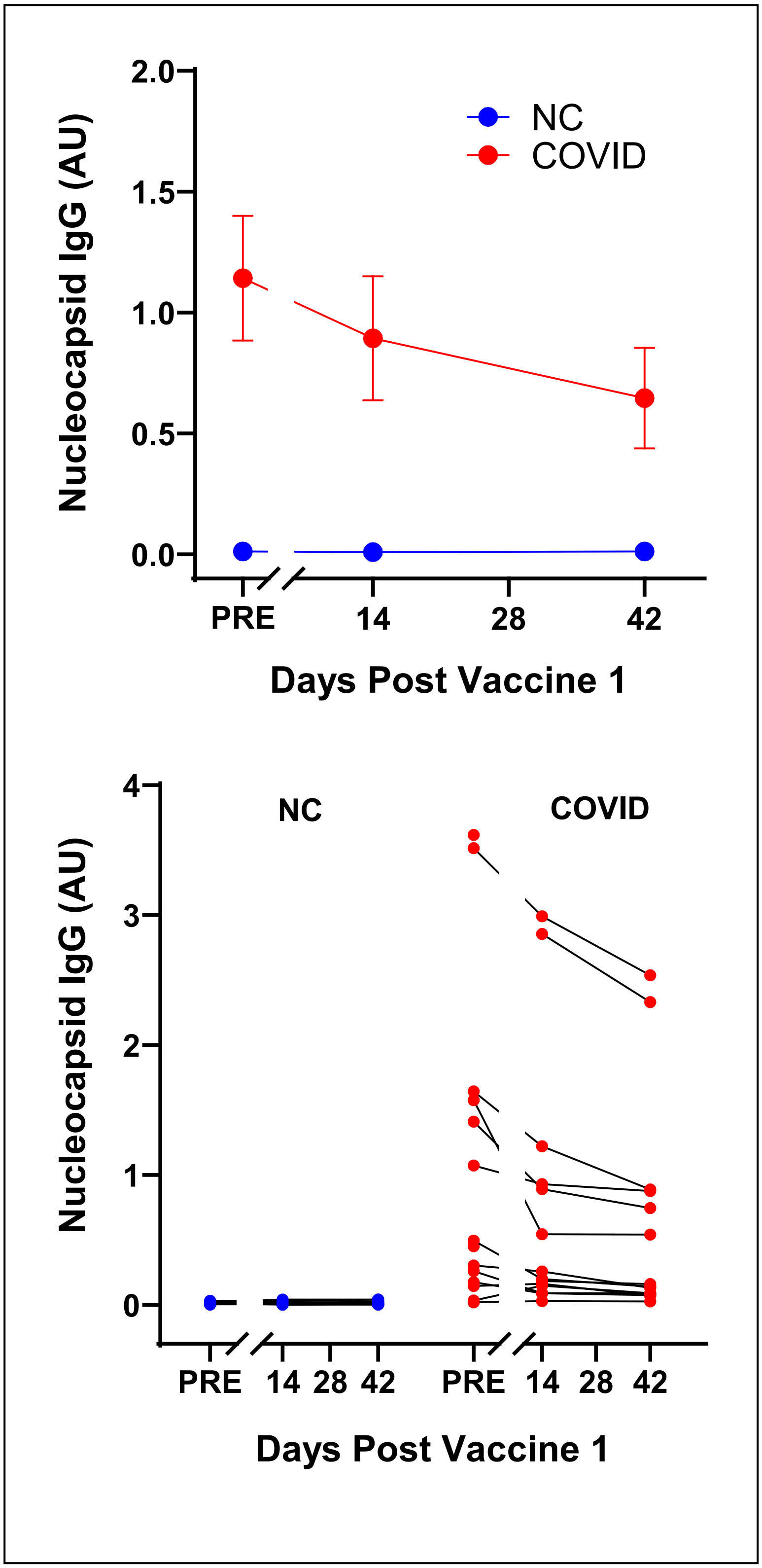

2.0
1.5
1.0
0.5
0.0
Nucleocapsid IgG (AU)
NC
COVID
PRE
14
28
42
Days Post Vaccine 1
4
3
2
1
0
Nucleocapsid IgG (AU)
NC
COVID
PRE
14
28
42
PRE
14
28
42
Days Post Vaccine 1

**Figure S3**

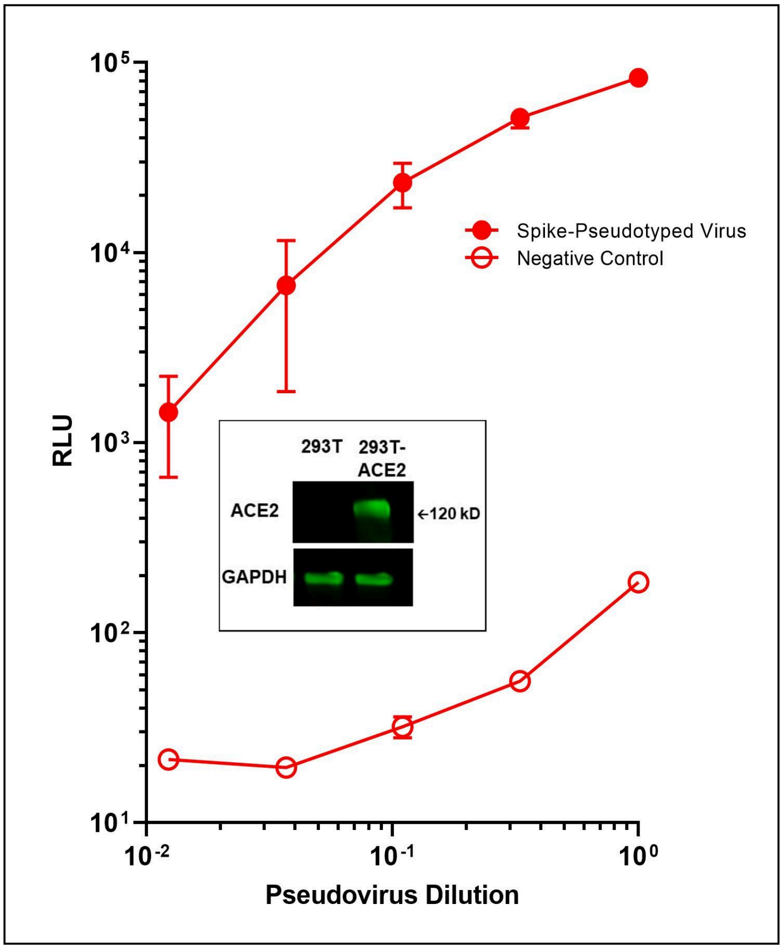

10^5
10^4
10^3
10^2
10^1
RLU
Spike-Pseudotyped Virus
Negative Control
293T
293T-ACE2
ACE2
←120 kD
GAPDH
10^-2
10^-1
10^0
Pseudovirus Dilution

**Figure S4**

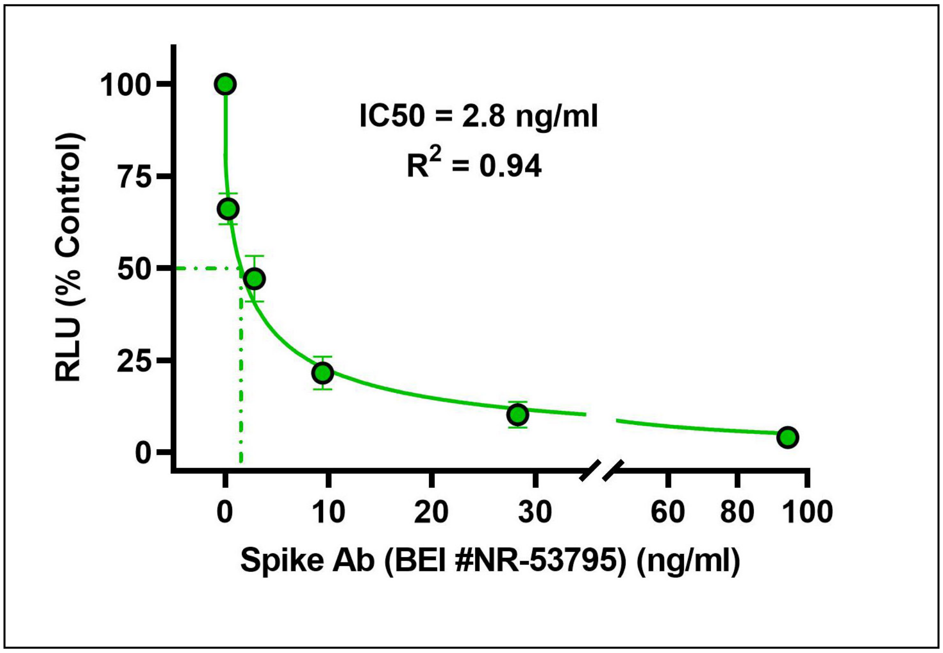
IC50 = 2.8 ng/ml
R2 = 0.94
RLU (% Control)
100
75
50
25
0
0
10
20
30
60
80
100
Spike Ab (BEI #NR-53795) (ng/ml)
